# An empirical Bayes framework for burden and dispersion association tests helps prioritize rare variants associated with Alzheimer’s disease

**DOI:** 10.64898/2026.06.15.26354742

**Authors:** Anjali Das, Chirag Lakhani, Vera M Mazeeva, Towfique Raj, David A Knowles

## Abstract

Rare genetic variants provide critical insight into the mechanisms underlying complex diseases, yet their study is limited by inherent statistical challenges, particularly in the noncoding genome where functional prioritization remains difficult. Here, we introduce `parmigiano`, an empirical Bayesian framework that systematically integrates functional annotations into existing rare variant association tests (RVATs), jointly learning annotation weights and a global variant filter threshold to enable trait-informed variant prioritization. We apply `parmigiano` to Alzheimer’s disease (AD) whole-genome sequencing data (12,900 cases and 23,846 controls) and perform both coding and noncoding RVATs, leveraging AD-relevant cell-type-specific predictions of variant regulatory effect. Integrating `parmigiano` significantly increases association yield across five existing RVATs, uncovering 23 candidate AD genes – 19 uniquely detected by our framework –including *SIGLEC10* and *HUNK*. Associations detected by `parmigiano` replicate more reliably in held-out data than those from the original RVATs and show higher overlap with known AD associations. `parmigiano` offers a unified, computationally efficient approach to variant prioritization, enabling scalable, interpretable rare variant analyses across coding and noncoding regions.

## 1 Introduction

Rare variants (RVs) are increasingly recognized as an important component of the genetic architecture of complex traits [1]. With the growing availability of whole-genome sequencing (WGS) data, RV analyses have become central in efforts to identify disease mechanisms beyond common-variant associations. In Alzheimer’s disease (AD), known loci explain only a modest fraction of the estimated genetic heritability, leaving a large portion of risk unexplained [2]. Large-scale WGS studies have shown that RVs contribute substantially to this gap across complex traits, with most signal arising from non-coding variants [3–5]. RVs often have larger per-allele effects due to negative selection [6–9] and vastly outnumber common variants, yet their contribution to disease remains difficult to characterize [10, 11]. Despite significant advances in the field, RV analyses face fundamental challenges related to statistical power, variant aggregation, and functional prioritization [12, 13].

Variant-level approaches, such as genome-wide association studies (GWAS), are highly effective for common variants but face challenges for RVs due to their sheer number and low minor allele frequencies (MAF) [14]. To address this, RVs are usually aggregated (e.g., by gene) to increase statistical power [15, 16]. Gene-level aggregation is relatively straightforward for coding variants, where boundaries are well defined and functional consequences are more predictable. As a result, most RV studies to date have focused on coding variation [17, 18].

However, this strategy overlooks the role of non-coding variation. Nearly 99% of the genome is non-coding, and most GWAS associations map to non-coding regions [19–21]. Extending RV analysis into the non-coding genome is therefore essential, but introduces significant methodological challenges. Existing approaches have defined non-coding testing units using fixed windows around transcription start sites, sliding windows, or individual *cis*-regulatory elements (CREs) [22–26]. Individual CREs are typically too small to provide adequate power, and sliding windows have a substantial multiple-testing burden. Results from these approaches are difficult to interpret since they do not resolve the causal gene. Aggregating variants in CREs predicted to regulate a specific gene can mitigate these issues, particularly when incorporating relevant cell-type specific information [27–30].

A potential concern with this strategy is the large variant set size per gene, especially as cohort sample sizes grow. Including large numbers of RVs per set can 1) increase inflation (false positives) in dispersion-based rare variant association tests (RVATs) [31], 2) substantially increase computational burden, and 3) add noise from uninformative variants (i.e., reduce statistical power), particularly in the non-coding genome where most variants are not functional. These problems will grow as WGS studies increase in size and discover more RVs, highlighting the need for principled approaches to leverage functional information to subset variants more likely to influence gene function, and thereby, disease.

In coding analyses, this is often achieved by restricting tests to predicted loss-of-function (LoF) variants, which enriches for large-effect alleles and substantially reduces the size of gene-level variant sets [32]. While effective, this strategy is inherently restrictive, as many pathogenic coding variants do not result in LoF. Moreover, there is no direct analogue of this approach for the non-coding genome, where regulatory variants span a wide range of functional impact and are distributed across diverse genomic contexts. Emerging variant effect predictors (VEPs), in particular cell-type–specific sequence-to-function models [33–37], provide rich prior information about non-coding activity. These predictions serve as proxies for intermediate molecular phenotypes such as gene expression, where population-based effect estimates (i.e., molecular quantitative trait loci, QTLs) are unavailable, for example for rare cell-states or variants. However, these annotations are heterogeneous, differ in their relevance across traits, and lack a clear binary interpretation analogous to LoF variation. Consequently, among the few existing RVATs that incorporate multiple VEPs, most either treat all annotations equally [38, 39] or apply ad hoc thresholds (e.g., PHRED-scaled CADD [40] > 5) to filter variants [41–43]. As a result, current approaches lack a systematic, trait-aware framework for optimally leveraging multiple VEPs to select likely functional non-coding variants within large testing regions, limiting both power and interpretability.

Here, we introduce Probabilistic Analysis of Rare variants Modeled In Genome-wIde ANnotation-informed Overdispersion test (`parmigiano`), an open-source framework (https://github.com/daklab/parmigiano) designed to systematically prioritize trait-relevant RVs for gene-based association testing. Like our previous model `gruyere` [29], parmigiano infers genome-wide annotation importance across coding and noncoding regions directly from the data in an empirical Bayes framework. `parmigiano` extends `gruyere` in three ways: 1) modeling variant effects as latent variables (random effects) with annotation-dependent priors, analogous to dispersion models such as SKAT [44]; 2) adaptively selecting functional variants in a model-based, trait-aware approach to improve power while reducing false positives and computational cost; and 3) integrating with five widely used RVATs: Burden [45], SKAT [44], SKAT-O [46], STAAR [38], and FST [39]. We inherit the ability of the underlying methods to handle related individuals through an observed relatedness matrix where applicable. Across simulations and in an application to the largest WGS dataset for AD to date, the Alzheimer’s Disease Sequencing Project (ADSP; 58K WGS release v5) [47, 48], we show that `parmigiano` increases the discovery and replication of AD-associated genes while substantially reducing the number of variants tested per gene. These results establish `parmigiano` as a scalable and principled foundation for RV analysis in large WGS studies.

## 2 Results

### 2.1 An empirical Bayes framework that augments existing RVATs

`parmigiano` is a hierarchical generalized linear mixed model (GLMM) framework for annotation-informed prioritization of trait-relevant variants in RVATs (Figure 1A). `parmigiano` models variant function as a weighted combination of multiple annotations *Z*, learning trait-specific annotation weights *τ* and a global filter threshold *T*, which together determine variant inclusion and weighting prior to downstream RVATs. The threshold *T* acts as a soft feature selection mechanism, pruning variants predicted to be neutral. This approach avoids fixed hard thresholds and flexibly integrates heterogeneous annotations, including deep learning–based and cell-type–specific predictions.

**Fig. 1.**
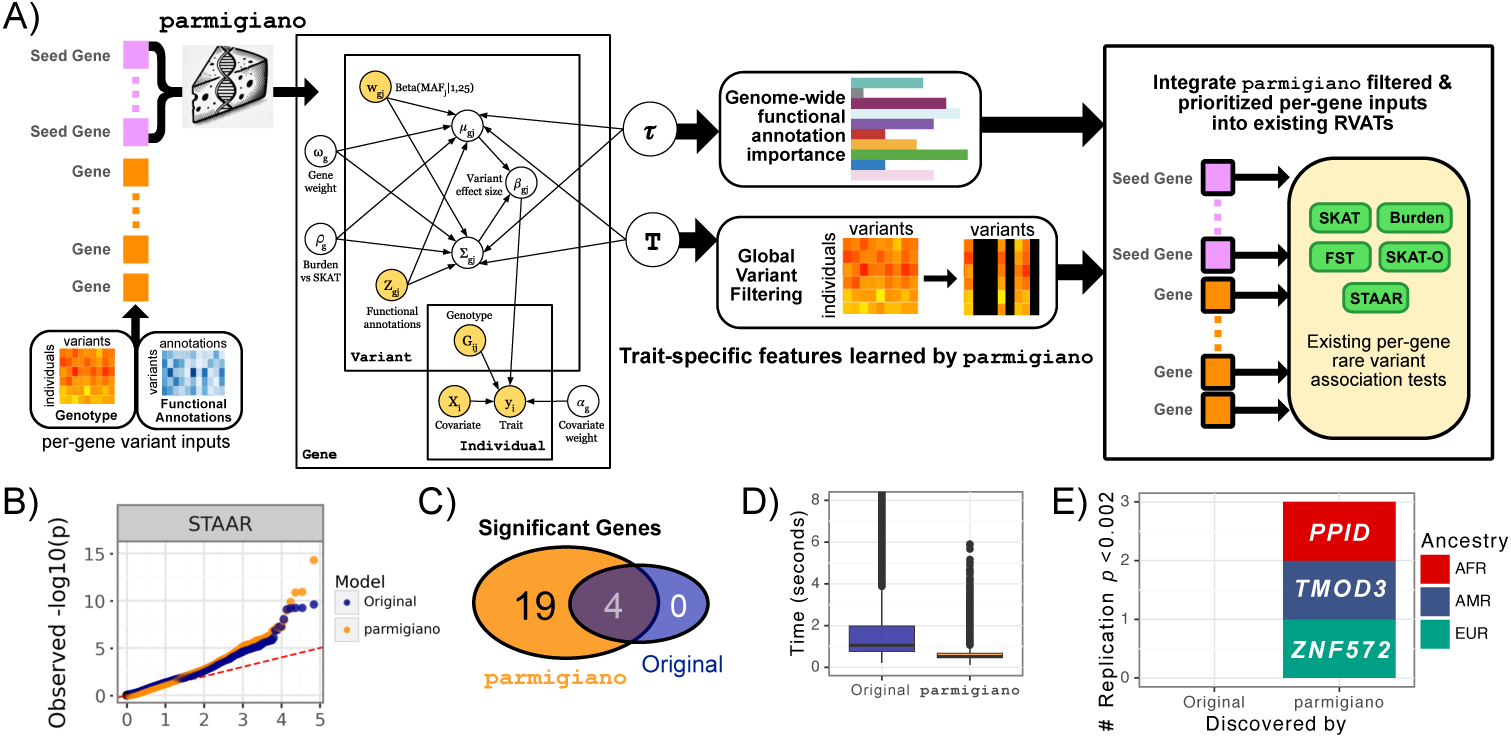
Variant prioritization with parmigiano enhances burden and dispersion association tests. **A)** Conceptual workflow. Seed genes (pink) are jointly fit in `parmigiano`. Plates denote dimensions (individuals, variants, genes). Filled circles indicate observed variables, and unfilled circles denote learned parameters. Arrows represent dependencies. `parmigiano` estimates annotation weights (*ω*) and a global variant filter threshold (*T*), which are applied genome-wide to prioritize variants for downstream RVATs. **B–E)** Downstream analyses integrating `parmigiano` with existing RVATs. **B)** Quantile–quantile plot of *p*-values comparing STAAR with its `parmigiano`-integrated counterpart. C) Overlap of significant genes identified across RVATs. **D)** Per-gene runtime (s) for STAAR before and after `parmigiano` integration. **E)** Number of discovery associations that replicate (*p <* 0.05*/*23), stratified by discovery type (original, `parmigiano`) and colored by ancestry group.

Similar to DeepRVAT [43] and our previous model `gruyere` [29], `parmigiano` is fit using a set of putatively trait-associated “seed” genes. The model is trained jointly across these genes within a scalable variational Bayesian framework. In DeepRVAT and gruyere, a variant’s effect is a learned deterministic function of its annotations, analogous to a functionally-informed burden test. In contrast, `parmigiano` is more flexible, modeling each variant effect as a latent variable whose *prior* (both mean and variance) depends on annotations through a learned function. This extension is equivalent to modeling *dispersion*. The globally-learned parameters (*τ*, *T*) are then applied genome-wide to prioritize variants in existing RVATs.

This strategy improves power by focusing on functionally relevant variation and reduces computational burden by excluding low-priority variants prior to testing. We first evaluate `parmigiano` in simulations to assess power and parameter recovery across varying heritability levels. We then apply `parmigiano` to the largest WGS dataset for AD to date, demonstrating improved gene discovery, replication, and computational efficiency.

### 2.2 Model performance on simulated data

#### Parameter recovery

We simulate data by sampling from the `parmigiano` model (with the priors specified in Methods 3.1) and using real ADSP WGS genotypes *G*, covariates *X*, and annotations *Z*. Across 500 simulations of 100 randomly selected genes, `parmigiano` accurately recovers genome-wide parameters. Learned and true annotation weights are highly correlated (*r̄*(*τ*) = 0.91), as are variant filter thresholds (r(*T*) = 0.75) (Figure 2A,B; Figure S1A). To assess the stability of parameter estimates, we perform 10 refits for a subset of 50 simulations. Parameter estimates are highly consistent across runs (Figure S1B), with low variability in global parameters (*σ̄*(*T*) = 0.012, *σ̄*(*τ_k_*) = 0.006). We recommend fitting `parmigiano` five times and averaging posterior estimates to further improve stability. We next evaluate performance across varying heritability levels. As expected, recovery of *ω_g_*, *ρ_g_*, and *τ* improves with increasing heritability (Figure S1C). Notably, under realistic heritability ranges (0.1–0.3), recovery of annotation weights and filter thresholds remains robust (r > 0.5; Figure S1D), while performance declines at very low heritability (< 0.1). Combined, these results indicate that `parmigiano` reliably learns trait-specific parameters under realistic genetic architectures.

**Fig. 2.**
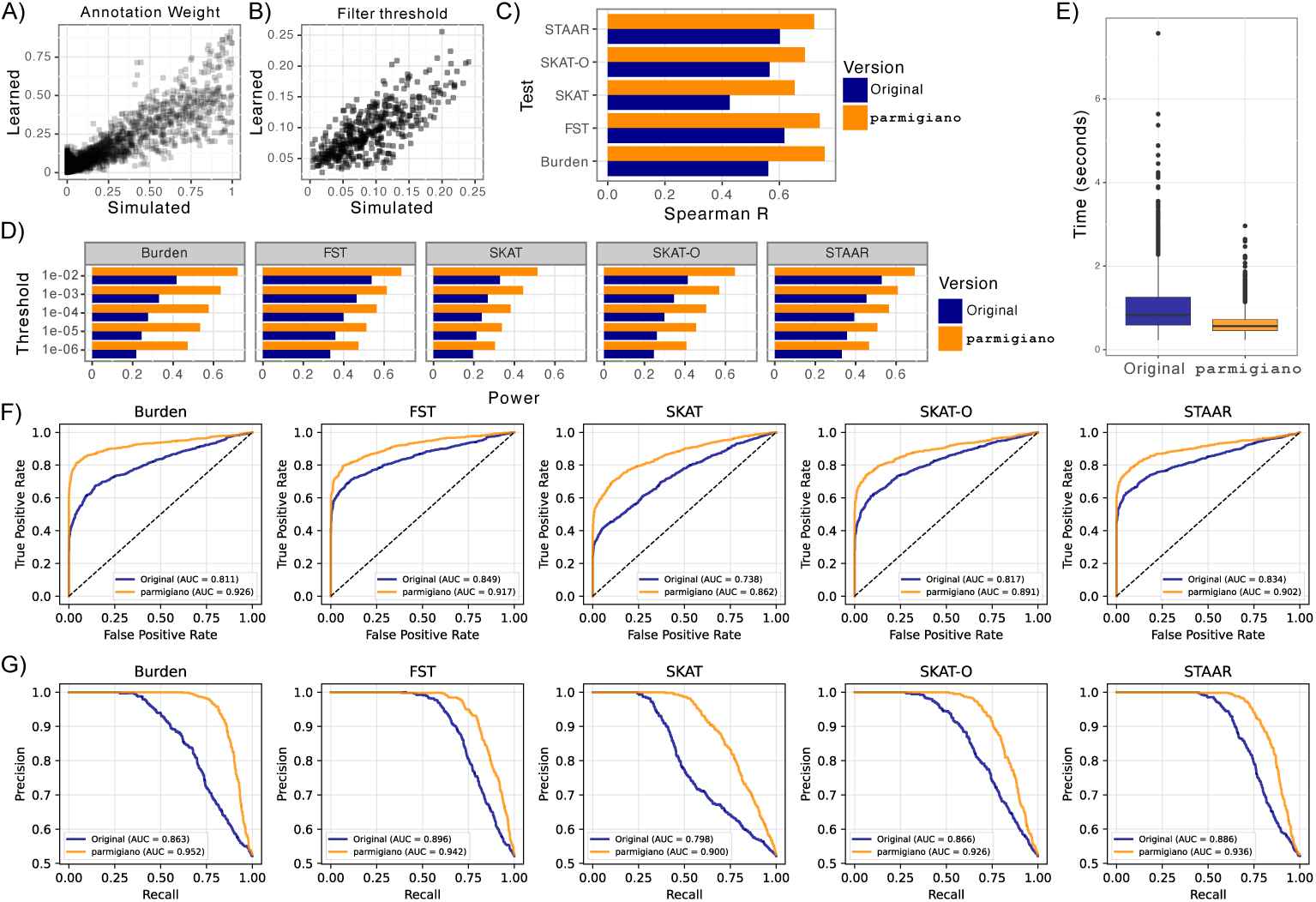
In simulations parmigiano recovers key parameters and improves power and computational efficiency across RVATs. **A)** Learned versus true annotation weights *ω* across 500 simulations. **B)** Learned versus true variant filter thresholds *T*. **C)** Spearman correlation between simulated gene weights *|ω_g_|* and *-* log_10_(*p*-values) across RVATs, comparing original and `parmigiano`-integrated tests. **D)** Power across *p*-value thresholds for original vs. `parmigiano`-integrated RVATs, defined as the proportion of genes with *p*-values ≤ a given threshold among genes with *|ω_g_| >* 1. **E)** Per-gene runtime under the FST framework for original vs. `parmigiano`-integrated tests. **F)** AUROC and **G)** AUPRC for distinguishing null versus non-null genes (*|ω_g_| >* 1) across RVATs, comparing original and `parmigiano`-integrated tests.

#### Power assessment

After confirming parameter recovery, we evaluate the impact of `parmigiano` on downstream RVAT performance. We simulate phenotypes that preserve case-control balance, covariate effects, and genetic structure observed in the ADSP data (Methods 3.1). Ground-truth gene weights (*ω_g_*) are defined per gene, with larger values indicating stronger effects of gene function on disease risk. We simulate data for 1000 null genes (*ω_g_* = 0) and 1000 non-null genes (*|ω_g_|* > 1).

We apply both standard and `parmigiano`-integrated RVATs and assess concordance between *|ω_g_|* and - log_10_(*p*-values). Correlations are consistently higher for `parmigiano`-integrated tests (Figure 2C), indicating improved prioritization of causal genes. Empirical power is also higher across *p*-value thresholds (Figure 2D). Separation between null and non-null genes is more pronounced for `parmigiano` (Figure 2F–G).

#### Runtime and scalability

We evaluate seed gene fitting time, scalability, and computational cost of the downstream RVAT. Fitting time increases with gene count, while annotation count has minimal impact (Figure S2A). Both memory usage and fit time scale linearly with sample size. Joint fitting converges in approximately one hour on CPU or 25 minutes on GPU using N = 50,000 samples, 100 seed genes, and 10 annotations, requiring roughly 20 GB of memory (Figure S2B). For very large datasets, subsampling 10,000 or more individuals yields comparable genome-wide estimates of *τ* and *T*.

After fitting, `parmigiano` is integrated into downstream RVATs, reducing computational cost due to the reduced number of prioritized variants to be tested. Because RVAT runtime scales with the number of variants per gene, applying the variant filter threshold decreases per-gene runtime in the FST framework from 1.06 to 0.63 seconds (Figure 2E). Despite the initial model-fitting step, this two-stage approach reduces overall runtime and improves scalability for large sequencing studies.

### 2.3 Application to ADSP WGS Data

We apply `parmigiano` to RVs in WGS data from ADSP release v5 [47], using N = 17,054 (5,515 AD cases and 11,539 controls) European-ancestry samples for discovery (Methods). We evaluate four groups of gene-level tests that leverage both coding and non-coding RVs. Each group includes coding variants (both nonsense and missense), intronic variants with SpliceAI scores > 0.1 [49], and RVs within cell-type–specific cis-regulatory elements (CREs) predicted by the Activity-by-Contact (ABC) [50] model for brain cell types: microglia, oligodendrocytes, astrocytes, and neurons. Because over 30% of predicted CREs are cell-type specific, analyses are performed separately per cell type, along with a coding-only test. This results in 27.9 million total annotated RVs and an average of 845 variants per gene set. We incorporate 14 functional annotation categories spanning coding, conservation, predicted pathogenicity, and deep learning–derived regulatory predictions (Methods 3.5.1).

### 2.4 parmigiano identifies deep learning-based annotations enriched for AD-associated variants

We fit `parmigiano` on ADSP WGS data using the 100 genes with strongest AD association according to STAAR as seed genes for each cell type (microglia, neurons, oligodendrocytes, astrocytes) and for a coding-only set. To avoid data leakage we use 5-fold cross-validation, fitting on a random 80% of seed genes per fold. Final *τ* and *T* estimates (Figure 3A) are averaged across all five folds and used for non-seed genes. For seed genes, weights are averaged only over the four folds in which the gene was held out. Learned *τ* and *T* estimates are stable across folds. We assess robustness to seed gene selection using five alternative gene sets, each defined as the top genes from different RVATs (Figure S3). Despite limited overlap among gene sets (Figure S3A), learned filter thresholds are highly consistent (range 0.13 - 0.15), and annotation weights vary minimally across fits (mean range 0.03; Figure S3B). This stability translates to highly consistent variant-level outputs: 88% of variants are classified identically across models (Figure S3E), variant weights are strongly correlated (mean Pearson’s R = 0.98; Figure S3C), and distributions are nearly identical (Figure S3D,F). Together, these results demonstrate `parmigiano’s` robustness to seed gene selection.

**Fig. 3.**
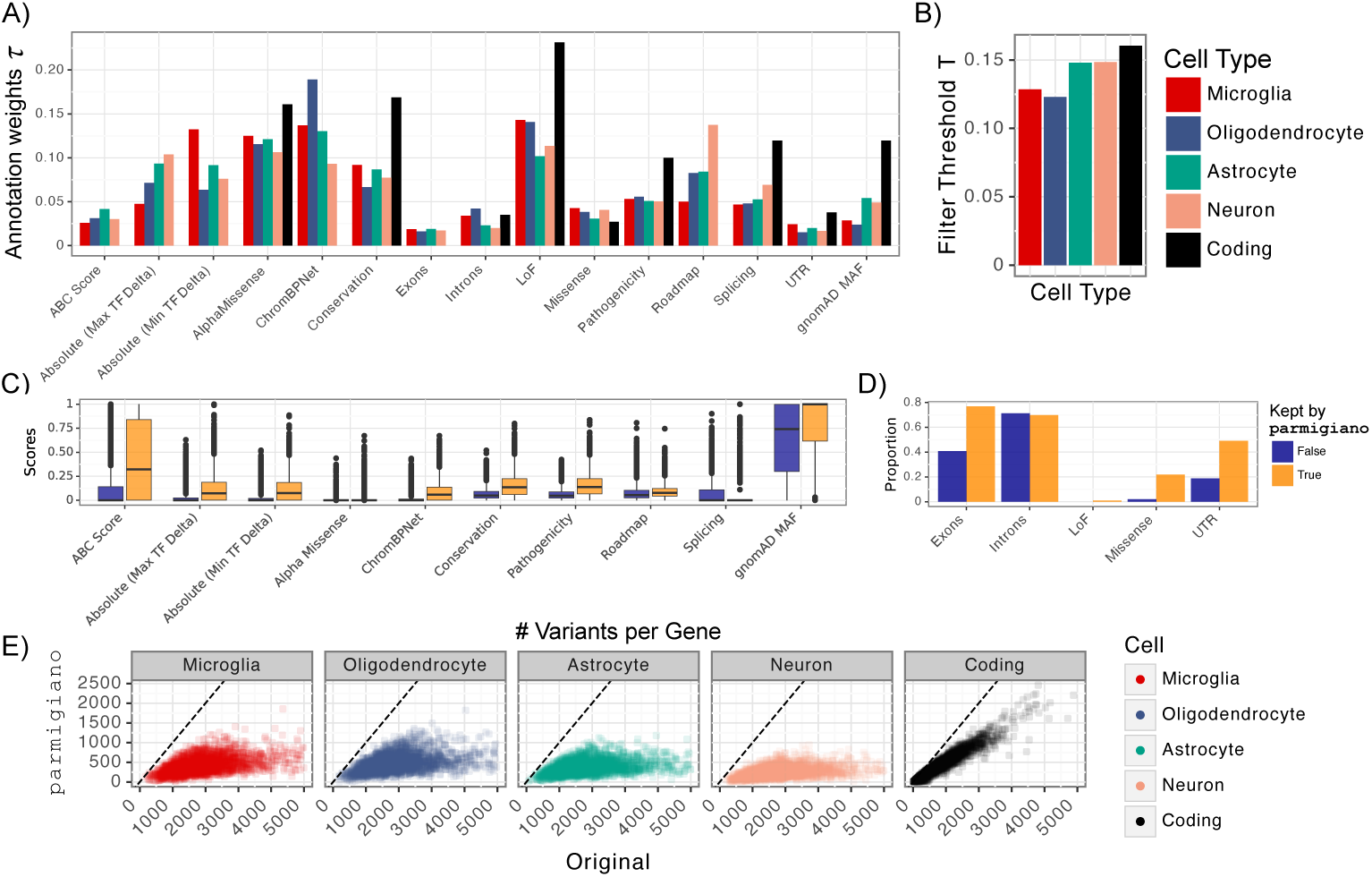
Variant prioritization learned by parmigiano. **A)** AD-specific annotation weights *ω*. **B)** Learned variant filter threshold *T* across cell types. **C)** Distribution of continuous microglia annotations stratified by `parmigiano` filtering. **D)** Enrichment of binary microglia annotations among retained versus filtered variants. **E)** Per-gene variant counts for `parmigiano`-integrated versus original tests; dashed line indicates *x* = *y*.

Learned annotation weights *τ* identify loss-of-function (LoF) variants as the strongest contributors to AD-associated RV signal in coding regions, consistent with their outsized impact on gene function [51]. Deep learning VEPs including ChromBPNet-predicted chromatin accessibility [34], Enformer-predicted transcription factor binding [33] and AlphaMissense-predicted pathogenicity [52], show strong enrichment in cell-type specific tests. Filter thresholds vary slightly across cell-type analyses (Figure 3B), but the number of retained variants per gene is broadly consistent (Figure 3E). `parmigiano` substantially reduces the total number of variants tested per gene, from an average of 374 to 190 in coding-only tests, and from 1,013 to 199 in combined coding and non-coding (cell-type-specific) tests, reflecting its ability to remove variants unlikely to contribute to disease. As expected, coding variants are preferentially retained relative to noncoding variants.

To characterize learned prioritization, we compare functional annotation distributions between retained and filtered variants in microglia (Figure 3C,D). Most annotations show clear separation between these groups, even for those with low weights. Notably, due to consistently high learned weights relative to T, no LoF variants are filtered.

### 2.5 parmigiano boosts association discovery across 5 RVATs in AD

We integrate AD-specific `parmigiano` variant weights (*τ* and *T*) into five established RVATs: Burden [45], FST [39], SKAT [44], SKAT-O [46], and STAAR [38]. We use related individuals where supported (Burden, SKAT, STAAR; N = 17, 054) and unrelated individuals for tests that do not accommodate relatedness (FST, SKAT-O; N = 14, 802). Across all RVATs, `parmigiano` increases power (Figure 4), yielding 23 significant genes in total, including four also identified in original tests (Figure 1C). `parmigiano` recovers all associations identified by the original tests, i.e., no signal is lost.

**Fig. 4.**
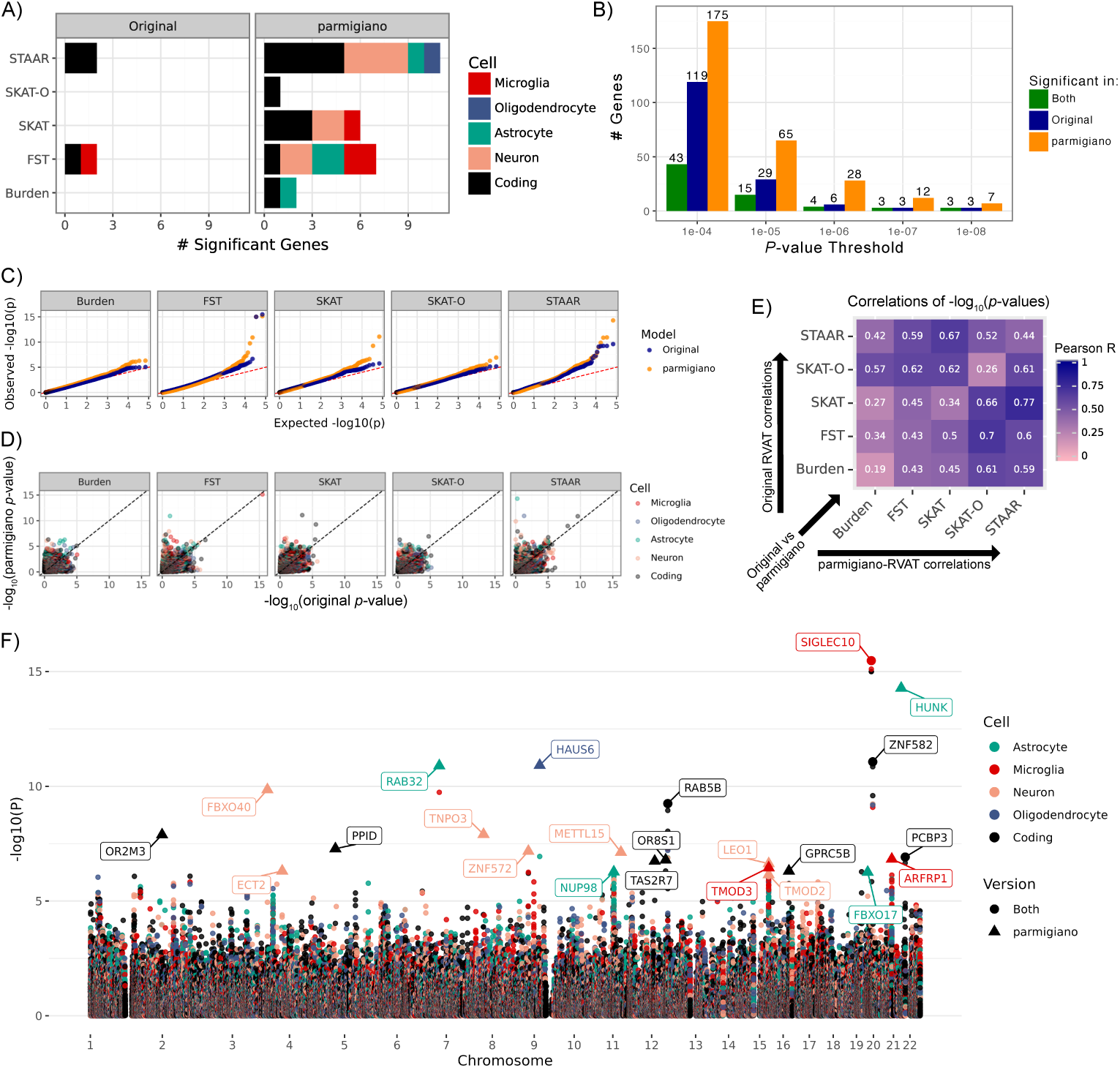
Original vs. parmigiano-integrated RVATs. **A)** Number of significant associations by RVAT colored by cell type. For genes significant in multiple cell types, the most significant cell type is shown. **B)** Number of significant genes identified across varying *p-*value thresholds for `parmigiano`, original, and both tests. **C)** QQ-plots of observed versus expected *-* log_10_(*p-*values) for original and `parmigiano`-integrated tests. **D)** Scatter plots of *-* log_10_(*p-*values) in `parmigiano`-integrated versus original tests across RVATs and colored by cell type. **E)** Pearson correlations of *-* log_10_(*p-*values) across tests. Upper-right: original; lower-left: `parmigiano`-integrated; diagonal: cross-version correlation for the same test. **F)** Manhattan plot across cell types and tests. The *Y* -axis shows *—* log_10_(*p*-value) for each gene and *X*-axis shows genomic position. Points are colored by cell type and shaped by which version (`parmigiano`, original, or both) the association is identified by. For genes significant across multiple cell types, the most significant association is labeled.

Significance thresholds are defined per cell type using Bonferroni correction across all genes and cell types (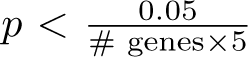, which is between 5.1 × 10*^-^*^7^ and 7.2 × 10*^-^*^7^ depending on the cell-type). Functionally uninformed tests (Burden, SKAT, SKAT-O) identify no significant associations in the original analyses, whereas `parmigiano`-augmented versions detect multiple significant genes (SKAT-O:1, Burden:2, SKAT:6). FST and STAAR find associations in their original forms but show substantial gains with `parmigiano`: STAAR increases from two (*RAB5B*, *ZNF582*) to eleven genes, and FST from two (*PCBP3*, *SIGLEC10*) to seven. QQ- and scatter plots demonstrate these improvements in power (Figure 4C,D). Across varying significance thresholds, `parmigiano` consistently identifies more associations (Figure 4B) without inflating the genomic control factor.

Correlations between original and `parmigiano`-integrated versions of the same test are modest (average R = 0.33), reflecting differences in underlying variant sets (Figure 4E). Concordance is lower for functionally uninformed tests (average R = 0.26) than for informed tests (average R = 0.44). Notably, cross-method correlations are higher for `parmigiano`-integrated analyses than for original tests, suggesting that `parmigiano` harmonizes signal detection across RVAT frameworks.

The strongest signal arises from the non-coding association observed at *SIGLEC10* in microglia, detected by both `parmigiano`-integrated and original RVATs (Figure 4F). `parmigiano` keeps 121 of the original 609 variants, with the signal driven primarily by LoF risk variants rs763006925 and rs201048913, and further boosted by the inclusion of microglia-specific ABC variants. *SIGLEC10* encodes a sialic acid–binding immunoglobulin-like lectin that is abundantly expressed in microglia and has been implicated in late-onset AD [53, 54]. A recent study shows increased *SIGLEC10* expression proximal to amyloid-*β* plaques and progressive upregulation in AD tauopa-thy. Transgenic expression of human *SIGLEC10* in microglia in a mouse model exacerbates both amyloid and tau pathology *in vivo*, effects that are reversible with anti-*SIGLEC10* antibody treatment [53]. Together, these findings provide biological support for our observed microglia-specific association at *SIGLEC10* and highlight its likely role in AD pathogenesis.

We identify *HUNK* (hormonally upregulated neu-associated kinase) in astrocytes as the most significant non-coding association uniquely detected by `parmigiano`, driven by `parmigiano`-STAAR (p = 5.2 × 10^-15^, Figure 5A). STAAR combines per-annotation burden, SKAT, and ACAT [55] tests via Cauchy combination, which we decompose to identify the driving annotation: the Enformer maximum TF delta annotation (Figure 5B). The original analysis includes 480 *HUNK* variants with MAC > 1, while `parmigiano` keeps 15 variants enriched in astrocyte-specific predicted CREs and largely excludes intronic variants (Figure 5C). These retained variants are generally rarer and exhibit greater case–control MAF differences than variants in the original analysis, unmasking the association that is otherwise diluted in the full variant set (Figure 5D). Three of the 15 `parmigiano` variants have TF delta scores greater than 0.1 (upper quartile), including two doubletons observed only in controls. Under the covariate-only null model, three carriers of these high TF delta variants are predicted to be AD cases (based on covariates) but are observed to be controls (two *APOE* -e4 homozygotes and one heterozygote), consistent with a potentially protective *HUNK* RV association (Figure 5E). Other carriers of these variants are already correctly predicted under the covariate-only null model and therefore contribute less to the association signal.

**Fig. 5.**
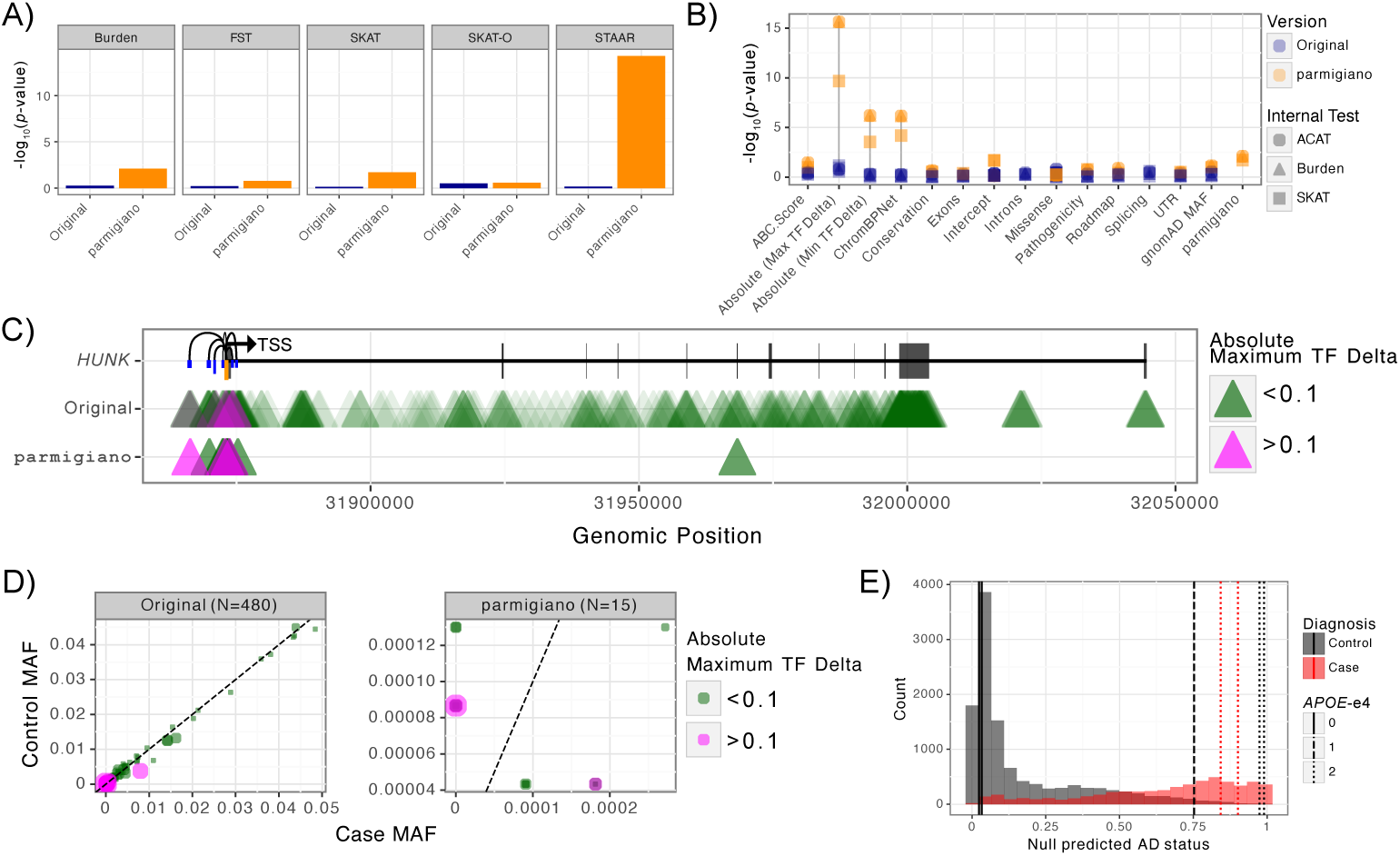
*HUNK* association in astrocytes identified by parmigiano-STAAR. **A)** *-log*_10_(*p-*values) across RVATs and by version. **B)** Internal STAAR *-log*_10_(*p-*values) across annotations and tests, with points shaped by test (ACAT-V, Burden, SKAT) and colored by version (original versus `parmigiano`. **C)** Genomic position of *HUNK* and variants included in the original vs. `parmigiano` analyses. Thin horizontal line shows gene start and end; black segments denote exonic regions; loops denote ABC-predicted regions (enhancers in blue, promoter in orange). Triangles denote variants filled by TF delta score. **D)** Control versus case MAF scatter plots for the original and `parmigiano` analyses (MAC *>* 1), with points colored and sized by TF delta score. **E)** Histogram of fitted AD predictions under null covariate model filled by true AD status. Vertical lines denote carriers of a high TF delta variant (*>* 0.1), with dash type indicating *APOE-e4* status.

*HUNK* is a serine/threonine kinase best characterized in breast cancer, where it has been implicated in progression and is highly expressed in HER2-positive breast cancer cell lines [56]. In the AD literature, *HUNK* has been reported as downregulated in a protein–protein interaction network derived from transcriptomic analyses [57]. Together, these observations are consistent with the protective direction of effect observed here, and provide a promising candidate for more systematic evaluation in AD cohorts.

We next compare the 23 significant gene associations to brain- and AD-relevant datasets. Three genes (*GPRC5B*, *TNPO3*, *RAB5B*) colocalize with brain-associated GWAS signals in recently published isoMiGA [58], BigBrain [59, 60], and SingleBrain [61] datasets (Figure S5A–C). Across varying significance thresholds, `parmigiano`-identified genes show consistently greater overlap with BigBrain and SingleBrain QTLs than genes identified by original RVATs (Figure S5E). `parmigiano`-significant genes also display higher expression across fourteen GTEx brain tissues [62] compared to non-significant genes (one-sided t-test, p = 4.3 × 10*^-^*^6^; Figure S5D). We find that across varying significance thresholds, `parmigiano`-identified genes are more enriched for Agora genes [63], a curated set of N = 947 genes associated with AD through various analyses, than original tests (Figure S5F). Combined, these results indicate that associations prioritized by `parmigiano` are more consistently supported by independent brain- and AD-relevant datasets than those identified by original RVATs alone.

### 2.6 parmigiano **enhances replication of RV associations**

We evaluate replication of significant associations from the European-ancestry discovery cohort in three independent validation datasets of European (EUR), African (AFR), and Hispanic (AMR) ancestries (Table 2; Methods 3.5). Associations identified by `parmigiano` show higher replication - log_10_(*p*-values) in EUR compared to those from original RVATs (one-sided t-test, p = 3.09 ×10*^-^*^5^; Figure 6A). This improvement is modest in AFR (p = 0.045) and not observed in AMR (p = 0.78), consistent with the ancestry specificity of RV associations [64].

**Fig. 6.**
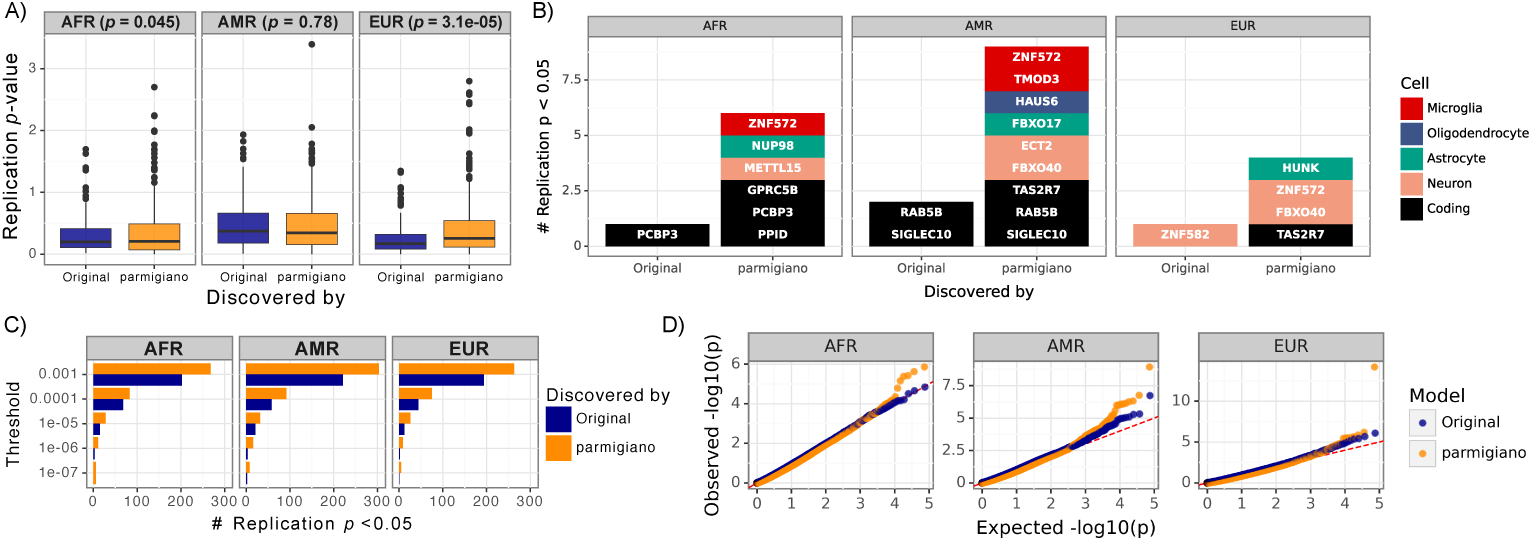
Increased replication of parmigiano-discovered genes across ancestry groups. **A)** Boxplot of *-log*_10_(replication *p*-value) for significant gene–cell associations identified in discovery split by discovery type (original vs. `parmigiano`) and ancestry. Each point represents a replication *p*-value; we show *p*-values from all RVATs (5 original and 5 `parmigiano`-integrated), for a total of 10 points per gene–cell association. **B)** Number of significant associations identified in discovery that nominally replicate (*p <* 0.05), split by discovery type (original, `parmigiano`) and ancestry and filled by cell type. We select the cell type with the smallest replication *p-*value for genes with multiple gene-cell discovery associations. **C)** Number of genes with replication *p <* 0.05 (x-axis) across varying discovery significance thresholds (y-axis) faceted by ancestry group and filled by discovery type. **D)** QQ-plots of observed versus expected *-log*_10_(*p*) for FST and `parmigiano`-FST for replication cohorts by ancestry.

Despite limited power for RV replication [17], nominal replication (p < 0.05) is observed for 15 of 23 `parmigiano`-identified genes and all four original associations in at least one ancestry (Figure 6B). Two of the three most significant discovery associations replicate: *SIGLEC10* in AMR (p = 0.028) and *HUNK* in EUR (p = 0.032). Although *HUNK* is astrocyte-specific in discovery, it also shows a strong microglial signal in AMR (p = 0.0013). After correcting for multiple testing (p < 0.05/23 = 0.0022), none of the original associations replicate, whereas three `parmigiano`-identified genes do (Figure 1E). All three—*PPID*, *TMOD3*, and *ZNF572* —have been previously implicated in AD [65–67].

Across a range of discovery thresholds, `parmigiano`-identified associations consistently show higher nominal replication rates than original RVATs across all ancestry groups (Figure 6C). Importantly, `parmigiano` weights generalize across datasets: using variant prioritization learned in discovery, `parmigiano`-integrated RVATs show higher power than unweighted tests in all replication cohorts (Figure 6D).

## 3 Methods

### 3.1 parmigiano **Model Framework**

`parmigiano` is a hierarchy of per-gene GLMMs (Figure 1A, Table 1)

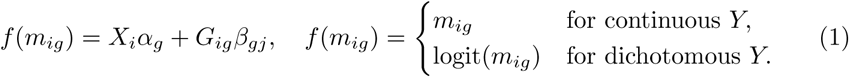

where E[*Y_ig_*] = *m_ig_* for individual *i* and gene *g*; *X_i_* = (*X_i_*_1_, …, *X_ic_*)*^T^* denote *c* covariates (e.g., age, sex, ancestral principal components, sequencing center); and *G_ig_* = (*G_ig_*_1_, …, *G_igp_*)*^T^* denote genotype dosages for p variants linked to gene *g*. Covariate weights *ς_g_* have prior,

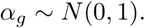

We use a Bernoulli likelihood for dichotomous traits and a Gaussian for continuous traits. Per-gene *g* variant *j* effect sizes *β_gj_* have prior

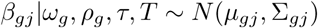

where

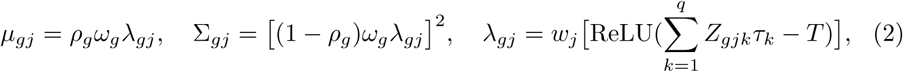

*ρ_g_* ∼ Beta(0.5, 0.5), *ω_g_ ∼* N(0, 1), *τ ∼* Dirichlet(**1***_q_*), *T* ∼ Beta(2, 20). The prior on variant effects *β_gj_* depends on the proportion of burden versus dispersion signal *ρ_g_*, learned gene weight *ω_g_*, transformed MAF w*_j_* = Beta(MAF*_j_|*1, 25), functional annotations *Z_gj_*, genome-wide annotation importance *τ* and variant filter *T*. The primary innovation of `parmigiano` is the construction of *β_gj_*, which (1) prioritizes functional annotations by a trait-specific global weight *τ*, (2) excludes variants with insufficient predicted functional relevance, using a learned threshold *T* on the weighted annotations 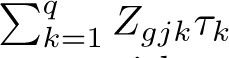, and (3) integrates (and weights) both burden and overdispersion components with *ρ_g_*.

**Table 1.** Summary of parmigiano Variables.

| Variable | Shape | Type | Description |
| --- | --- | --- | --- |
| $Y$ | $n \times 1$ | Observed | Phenotypes for $n$ samples |
| $X$ | $n \times c$ | Observed | $c$ Covariates for $n$ samples |
| $G_g$ | $n \times p_g$ | Observed | Genotypes for $p$ variants in gene $g$ |
| $Z_g$ | $p_g \times q$ | Observed | $q$ Functional annotations per variant |
| $w_j$ | $p_g \times 1$ | Observed | Transformed MAF weights |
| $\alpha_g$ | $c \times 1$ | Learned | $c$ Covariate coefficients |
| $\beta_g$ | $p_g \times 1$ | Learned | Variant effect sizes for $p$ variants |
| $\rho_g$ | $1 \times 1$ | Learned | Proportion burden versus dispersion |
| $\omega_g$ | $1 \times 1$ | Learned | Gene importance weights |
| $T$ | $1 \times 1$ | Learned | Variant filter threshold |
| $\tau$ | $q \times 1$ | Learned | Functional annotation weight |

#### Generating synthetic phenotypes

To assess parameter recovery, we simulate phenotypes *Y* by sampling directly from Equation 1 using the priors specified above, while retaining real ADSP WGS genotypes, covariates, and annotations.

To assess power, we simulate phenotypes designed to preserve the genetic and covariate structure of AD WGS data. We first fit `parmigiano` on real data to obtain empirical parameter distributions, using global parameters *τ* and *T* estimated from the top 100 microglia STAAR genes and fixed across all simulations. We simulate phenotypes for 2000 randomly selected genes, retaining real AD WGS genotypes G*_ig_* and covariates X*_i_* while sampling gene-specific parameters *ω_g_* and *ρ_g_* from their empirical distributions to generate phenotypes according to Equation 1. This design preserves the genetic architecture and covariate structure of the real data while letting us control the ground truth gene-level effects.

Of the 2000 genes, 1000 are set to null (*ω_g_* = 0) and 1000 to non-null. In non-null simulations, *ω_g_* was sampled from N(0, 1) (as in the generative model in Equation 1); samples yielding *|ω_g_|* < 1 are resampled to enforce separation between null and non-null effect sizes. Standard and `parmigiano`-integrated RVATs were applied to each phenotype, and resulting *p*-values were compared to ground-truth *ω_g_* to assess power.

### 3.2 Relation to other RV association tests

Several established burden and dispersion tests arise as special cases of `parmigiano`.

#### Burden Test

In standard burden tests, all variants *j* in gene *g* are assumed to have the same effect [45]. When *ρ_g_* = 1 then C*_gj_* = 0, `parmigiano` reduces to a functionally-informed burden test where variant effects are fully determined by the annotations, since then

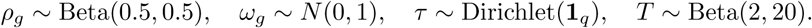

equivalent to our previously proposed `gruyere` model [29]. This reduces to a standard burden test if there are no annotations (or if model fitting chooses to give them no weight, i.e., *τ_k_* = 0 for k ≠ 0) and the filter threshold does not exclude any variants (*T* = 0).

#### Dispersion Test

When *ρ_g_* = 0, our model is a variance-component (overdispersion) test, which assesses whether variants *j* in gene *g* contribute to the *variability* in a trait, where *ρ_g_* controls the strength of dependence on annotations. This variability is modeled by Σ*_gj_* in `parmigiano`, since

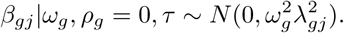

The dependence on annotations is analogous to FST (although they do not learn genome-wide weights, or perform model-based variant filtering). Without annotations and variant filtering, we have equivalence to SKAT [44].

#### Unified Test

Analogously to SKAT-O, we combine signals from burden- and dispersion-based tests by learning *ρ_g_*, which models the correlation between variant effect sizes within a gene, since

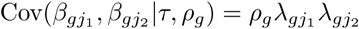

after integrating out *ω_g_*. For large *ρ ≈* 1, variants in the same gene with the similar λ (e.g. because they have similar annotations *Z*) will have similar *β* since then *β_gj_ ≈ ω_g_λ_gj_*. For small *ρ* variants are uncorrelated (but variants with larger λ have larger effects a priori).

### 3.3 Model fitting

Learning functional annotation weights *τ* and filter threshold *T* from thousands of genes genome-wide is both computationally demanding because of the large number of genes and especially RVs involved, and analytically unnecessary, as only the fraction of genes associated with trait *Y* will provide meaningful information about which annotations are important. Therefore, we fit *τ* and *T* from a broad subset of potentially disease-relevant genes. Our `parmigiano` pipeline provides a workflow that uses conventional RVATs to identify seed genes (following DeepRVAT [43]). We tested sensitivity of `parmigiano` to the set of seed genes provided and find them to be robust. For the final results we select the top 100 associations from STAAR.

We find that naive variational inference for the model can struggle to effectively learn the variance Σ*_gj_* but significantly improves when we use the “non-centered parameterization” [68],

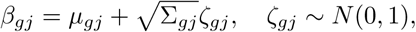

so the variational distribution is on ζ rather than *β* directly. We select a Dirichlet prior for the annotation weights *τ* to ensure identifiability between *τ* and *ω_g_*. Without constraining *τ* to a fixed sum, we can switch *ω_g_* for *ω_g_*/c and *τ* for c*τ* for any positive constant c without changing the likelihood, leading to non-identifiability between gene and annotation weights.

We approximate the true posterior distribution by maximizing the Evidence Lower Bound (ELBO) (equivalently minimizing the Kullback-Leibler (KL) divergence) [69] with SVI using the pyro probabilistic programming language [70, 71]. The posterior distribution of latent variables *ς_g_*, *ω_g_*, *ρ_g_*, and *↼_gj_* is approximated with mean-field (i.e., diagonal covariance) normal distributions, while the parameters *τ* and *T* are optimized (achieved in pyro via a delta variational posterior). We use a learning rate of 0.1 with the Adam optimizer, train for 300 epochs, and draw 100 samples from the posterior to estimate the mean and standard deviation of the mean-field normal approximation.

Sensible initializations can help ensure consistent convergence of probabilistic models. Therefore, we optionally initialize parameters *τ* and *ω_g_* using a simpler burden-only model (*ρ_g_* = 1). If no initialization is applied, we recommend refitting `parmigiano` five times and averaging posterior results to ensure consistency in learned parameters.

### 3.4 Integrating globally-learned parmigiano parameters into existing RVATs

Once trait-specific estimates are fit on seed genes, they are applied genome-wide and integrated into existing RVATs. For each variant, we compute a score *λ_gj_* (Equation 2), which applies a threshold *T* to the annotation-weighted sum *Zτ*. Variants with *λ_gj_* = 0 are excluded from downstream analyses.

For methods that do not natively incorporate functional annotations (e.g., burden, SKAT, SKAT-O), variants are re-weighted by *λ_gj_*. For annotation-informed RVATs (e.g., STAAR and FST), variants are filtered identically, and *λ_gj_* is included as an additional annotation alongside existing functional annotations.

We implement `parmigiano` with five RVATs: Burden [45], SKAT [44], SKAT-O [46], FST [39], and STAAR [38]. Burden, SKAT, and STAAR are run using the STAAR R package, which allows incorporation of a genetic relatedness matrix (GRM). SKAT-O and FST are run using the FSTpackage, which does not accommodate related samples; these analyses are performed on unrelated individuals only.

The effect of individual annotations is tested separately in annotation-informed RVATs (e.g., STAAR and FST). For these tests, the inclusion of singleton variants can induce spurious associations when a binary annotation (e.g., LoF) contains only a single variant within a gene, allowing that variant to dominate the test statistic. To mitigate this effect, we include singleton variants only in the intercept term and apply functional annotations exclusively to variants with minor allele count (MAC) > 1. Annotations with zero variance within a gene-set are removed for that gene prior to testing.

### 3.5 Data overview: Applications to AD

#### WGS and Covariate Data

We analyze WGS and clinical data from the Alzheimer’s Disease Sequencing Project (ADSP v5) [47], comprising 36,746 individuals after quality control (12,900 AD cases, 23,846 controls). AD cases are defined as individuals with a reported diagnosis and age > 50 years, and controls are restricted to age > 65 years to reduce misclassification due to preclinical disease. During QC, we exclude the EFIGA cohort (N = 4, 086) due to systematic inflation in RVATs. Standard QC removes duplicate and technical replicate samples, prioritizes family-study phenotypes when overlapping with case–control studies, and excludes individuals with high missingness (> 10%) and variants with low genotyping rate (< 90%). Missing genotypes are mean-imputed.

Ancestry is inferred with Somalier [72] using 1000 Genomes [73], resulting in predominantly European (EUR, N = 21, 900), African (AFR, N = 5, 356), and Hispanic (AMR, N = 5, 294) samples. RVATs are performed separately by ancestry to account for ancestry-specific effects. For discovery, we use the European samples excluding the largest cohort (NIA Alzheimer Disease Centers, N = 4, 846), which is reserved for replication alongside AFR and AMR datasets (Table 2). Samples of East Asian (EAS) and South Asian (SAS) ancestry were excluded from downstream RVATs analyses due to limited sample size and power but are included in the total QC’d sample count.

**Table 2.** Overview of discovery and replication datasets.

| Dataset | Ancestry | Sample size | Cases | Controls |
| --- | --- | --- | --- | --- |
| Discovery | EUR | 17,054 | 5,515 | 11,539 |
| Replication | EUR | 4,846 | 2,994 | 1,852 |
| Replication | AFR | 5,356 | 1,761 | 3,595 |
| Replication | AMR | 5,294 | 1,508 | 3,786 |

To account for population structure, we include genotype principal components (PCs). When including related individual we use the empirical kinship matrix. Of 36,746 individuals, 31,543 (11,227 cases, 20,316 controls) are considered unrelated by KING [74]. We apply the default settings of the FastSparseGRM pipeline [75] to determine unrelated samples, ancestry PCs, and a block-diagonal sparse ancestry-adjusted genetic relatedness matrix (GRM). Covariates included in all analyses are sex, age, *APOE-e4* and *APOE-e2* genotypes, 20 ancestry PCs, a common variant polygenic risk score computed using PRS-CS on ADSP samples [76], sequencing platform, sequencing center, cohort, age^2^, age × sex, age^2^ × sex, and an intercept term.

#### Construction of Variant Sets

We define gene-level variant sets to assess the contribution of both coding and noncoding RVs (MAF ↘ 0.05) to AD risk. Coding variants are restricted to RVs within exonic regions, annotated using GENCODE [77]. Non-coding variants include (i) RVs within cell type–specific cis-regulatory elements (CREs) predicted by the Activity-by-Contact (ABC) model [50] for AD-relevant cell types (microglia, oligodendrocytes, astrocytes, neurons) and (ii) intronic RVs with nonzero SpliceAI-predicted splicing effects [49]. Nonzero SpliceAI intronic variants are included because our prior findings indicate that AD-associated RVs are enriched for SpliceAI predictions [29].

We calculate enhancer–gene connectivity using publicly available ATAC-seq and H3K27ac ChIP-seq data [27, 78] and apply the ABC model following standard guidelines [50] using hg38-aligned data. Candidate enhancer regions are called from ATAC-seq peaks using MACS2, enhancer activity is quantified as the geometric mean of ATAC-seq and H3K27ac ChIP-seq signal, and ABC scores are computed with cell-type averaged Hi-C data. For each gene and cell type, all elements E with ABC scores ≃ 0.02 are aggregated, and variants within these regions are assigned to the corresponding gene(s). For each cell-type-specific test, gene sets combine coding RVs, nonzero SpliceAI intronic RVs, and RVs within predicted CREs for that cell type. Each cell type, and the coding-only variant sets, are analyzed independently, resulting in five sets of results.

#### 3.5.1 Functional Annotations

We consider fourteen functional annotation groups: coding context (missense, intron, exon, UTR), loss-of-function (LoF) [51], splicing [49], conservation [79], pathogenicity [80, 81], brain-specific epigenetic marks from Roadmap [82], deep learning–based protein function disruption scores (AlphaMissense [52]), population-specific MAF [83], ABC scores [50], and cell-type–specific VEPs from deep learning models (Enformer [33] for TF binding; ChromBPNet [34] for chromatin accessibility). Continuous annotations are first PHRED-scaled following STAAR [38] and subsequently min–max scaled to the range [0,1] to ensure comparability when using multiple annotations in `parmigiano`. Binary annotations are encoded as 0/1 indicators. Detailed annotation construction, grouping, and preprocessing are described in Supplemental Methods 4.9 and Table S1.

## 4 Discussion

Here, we present `parmigiano`, a scalable framework for systematically integrating trait-relevant functional annotations into RVATs. `parmigiano` improves statistical power across five existing RVATs while reducing their computational burden. In analyses of the largest AD WGS dataset to date, `parmigiano` identifies associations with higher replication rates across independent validation cohorts and greater overlap with known AD- and brain-relevant datasets, supporting the biological relevance of the identified signals. As WGS datasets continue to expand and functional annotations become increasingly refined, `parmigiano` provides a robust approach for uncovering trait-specific RV associations.

Beyond improving the performance of existing RVATs, `parmigiano` functions as a general preprocessing framework for complex trait analyses. By explicitly learning which functional annotations are most informative for a given trait, the model provides insight into disease-relevant regulatory mechanisms. In AD, we observe strong enrichment for deep learning–based variant effect predictions, including transcription factor binding, chromatin accessibility, and pathogenicity, which remain underuti-lized. Enrichment of cell-type–specific annotations adds further biological resolution: by identifying which cell types drive association signals, `parmigiano` can help pinpoint both the relevant cell type(s) and target gene. Given the complex etiology of AD, risk genes may act in multiple cell types. This resolution will improve as cell-type–specific variant effect prediction models become more comprehensive and accurate.

While the current implementation of `parmigiano` requires individual-level WGS and functional annotation data as input, our held-out replication results suggest that learned weights generalize beyond the cohort in which they were derived. A key future direction is the direct application of `parmigiano` weights to summary-level data [84]. Although model training currently requires individual-level information, the resulting weights can in principle be applied to summary statistics, substantially broadening the range of datasets amenable to this approach.

In this study, we restrict annotation scores to be non-negative, treating them as measures of functional relevance rather than effect direction. While appropriate for variant prioritization, modeling effect direction is important for biological interpretation and could potentially further increase power. Although dispersion-based tests allow variants with opposing effects to contribute at the gene level, future extensions could incorporate directional predictions directly, enabling more precise prioritization of variants that increase or decrease disease risk.

A limitation of our ancestry-stratified approach is reduced power to detect associations shared across populations. While our focus on European ancestry reflects current sample size availability, some signals may only emerge in combined analyses. For example, *TREM2*, a well-established AD risk gene [85, 86], does not reach significance in individual ancestry analyses but approaches genome-wide significance when all ancestries are combined (p = 2.8 × 10*^-^*^6^). This highlights a key trade-off: ancestry-stratified analyses capture population-specific effects, while combined analyses improve power for shared signals. As more diverse cohorts become available, methods that jointly model shared and ancestry-specific rare variant effects will be increasingly important.

As WGS datasets expand, the number of variants per gene—particularly in noncoding regions—increases substantially. Consistent with prior work [31], we observe that SKAT-based methods can be sensitive to variant count, leading to inflation or deflation of *p*-values. By filtering variants unlikely to contribute meaningful signal, `parmigiano` reduces the number of variants per test and mitigates this effect (Figure S4). However, some sensitivity remains even after filtering, pointing to limitations in existing RVAT frameworks and the need for methods that better scale to large non-coding variant sets.

Finally, our analysis highlights both the benefit and complexity of incorporating cell-type–specific regulatory annotations into RV studies. We analyze each cell type independently, selecting the most significant association when multiple exceed significance thresholds. Because many predicted regulatory elements are cell-type specific, this approach avoids assumptions about shared regulatory architecture. Methods that explicitly model shared and cell-type–specific effects may further improve resolution and represent an important direction for future work.

In conclusion, `parmigiano` offers a unified framework for annotation-informed RV analysis across coding and non-coding regions. By improving power, interpretability, and scalability, `parmigiano` is well positioned to enable discovery as WGS studies expand and functional annotations continue to advance.

**Supplementary information.** Supplementary Figures 1–5, Supplementary Tables 1–2, Supplementary Methods and Supplementary Results.

## Data Availability

This paper uses the Alzheimers Disease Sequence Project Release 5 Whole Genome Sequencing data and Alzheimers Disease phenotype data.

## Acknowledgements

This work was supported by the Alzheimer’s Disease Sequencing Project of the National Institutes of Health under award number U01 AG068880-02. The content is solely the responsibility of the authors and does not necessarily represent the official views of the National Institutes of Health.

## Declarations

### 4.3 Ethics approval and consent to participate

Ethics approval and consent to participate were obtained through the original ADSP studies. Data access was granted through dbGaP (phs000572).

### 4.4 Consent for publication

Not applicable

### 4.5 Data availability

This paper uses the Alzheimer’s Disease Sequence Project Release 5 Whole Genome Sequencing data and Alzheimer’s Disease phenotype data. Functional annotations used in this study are publicly available; sources are described in the Supplementary Methods. Per-gene association results for all cell-type-specific tests, `parmigiano`-integrated and original RVAT results are provided as Supplementary Data.

### 4.6 Materials availability

Not applicable

### 4.7 Code availability

Code and instructions for running `parmigiano` is available on GitHub: https://github.com/daklab/parmigiano

### 4.8 Author contribution

A.D. and D.A.K. developed the methods. A.D. and D.A.K. wrote the manuscript. A.D. wrote the software code and performed the primary analyses. C.L. helped with data QC, preprocessing, and annotating variants. V.M.M. helped test software. T.R. gave input on AD relevance. All authors read, reviewed, and approved the final manuscript.

