## Supplemental Material for "An empirical Bayes framework for burden and dispersion association tests helps prioritize rare variants associated with Alzheimer’s disease"

### Supplemental Figures and Results

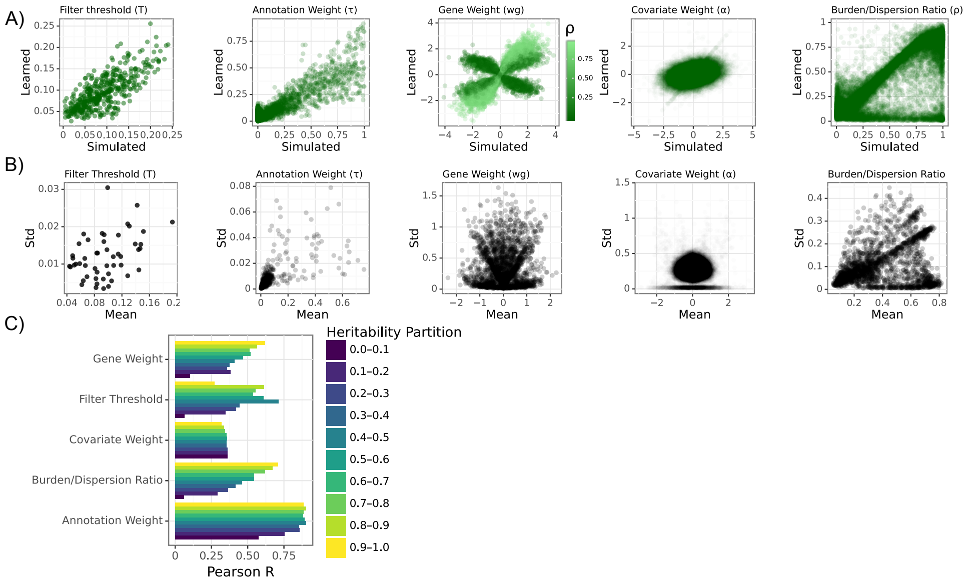


**Fig. S1 Simulation performance. A)** Learned versus true `parmigiano` parameters across 500 simulations. Gene weight $\omega_g$ is colored by simulated $\rho$. **B)** Standard deviation versus mean `parmigiano` parameters across 10 refits of 50 simulations. **C)** Pearson R correlations of parameter performance partitioned by genetic heritability bins.

We find that `parmigiano` accurately recovers all simulated parameters, with average Pearson correlations between learned and true weights across simulations: $\bar{r}(\tau) = 0.91$, $r(T) = 0.75$, $\bar{r}(|\omega_g|) = 0.62$, $\bar{r}(\rho_g) = 0.79$, and $\bar{r}(\alpha_g) = 0.53$ ( S1A). The scatter plot of learned versus true gene weights exhibits a cross-shaped pattern, indicating that while the magnitude of $\omega_g$ is learned accurately, the sign is sometimes swapped. This occurs when $\rho_g$ is close to 0, as the variance $\Sigma_{gj}$ depends on $|\omega_g|$ and therefore cannot distinguish between positive and negative effects. Because the magnitude is correctly recovered, this sign ambiguity is not a concern. In some simulations, $\rho$ is set close to 1 (burden) but is learned near 0 (dispersion). We do not observe the inverse scenario, which is reassuring, as dispersion captures burden signal but not vice versa. In these cases, we also observe higher standard deviations of $\rho$ across refits of the same simulation, indicating, as expected, that the model is uncertain about the exact value of $\rho$.

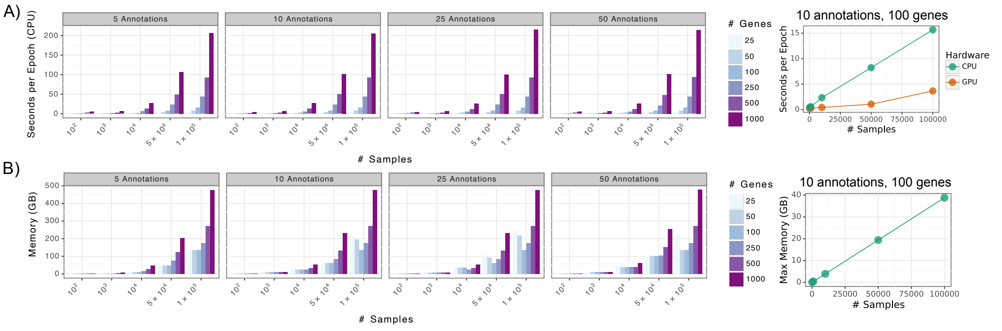


**Fig. S2 Time and memory in simulations. A)** Left: Seconds per epoch to fit `parmigiano` across varying number of annotations, sample sizes, and genes. Right: Time per epoch for fixed annotation (10) and gene (100) counts across varying sample sizes for CPU versus GPU. **B)** Left: Required memory (GB) to fit `parmigiano` across varying number of annotations, sample sizes, and genes. Right: Memory for fixed annotation (10) and gene (100) counts across varying sample sizes. **C)** Left: Scatter plot of time (seconds) to score a gene versus number of variants per gene for Original model versus `parmigiano` model within the STAAR framework. Right: Boxplot of original versus `parmigiano` time to score a gene within the STAAR framework.

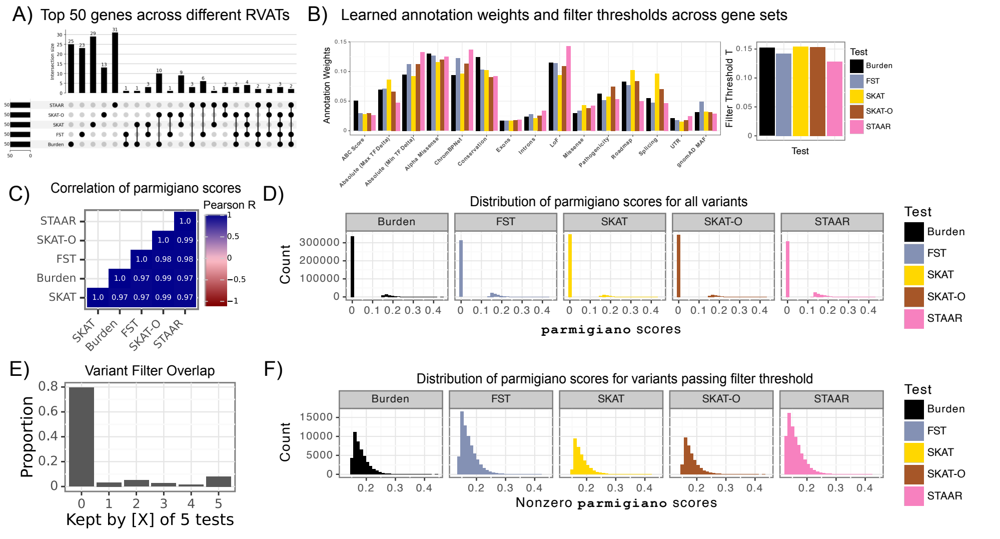


**Fig. S3 $\tau$ and $T$ sensitivity across varying seed genes (microglia). A)** Upset plot of overlap across top 50 genes defined by different RVATs (5 sets of seed genes). **B)** Learned $\tau$ and $T$ (in legend) across different seed gene sets. **C)** Correlation of variant weights $Z\tau$ across gene sets. **D)** Histograms of variant weights $Z\tau$ across 5 gene sets. **E)** Proportion of variant retained overlap across 5 sets of seed genes. Kept by 0 of 5 tests $\implies$ all five gene sets filter the same variant. **F)** Histograms of variant weights $ReLU(Z\tau - T)$ for non-filtered ie. non-zero variants across varying seed genes.

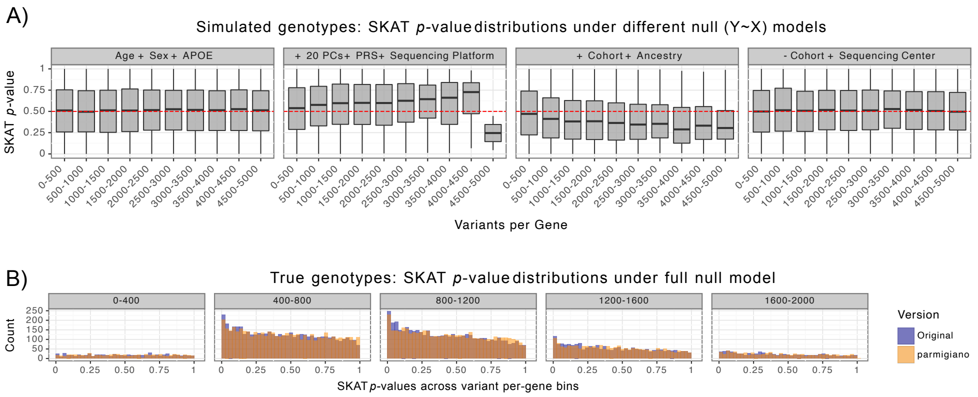


**Fig. S4 Calibration of SKAT $p$-values. A)** SKAT $p$-values across different null models, segmented by number of variants per gene. **B)** SKAT $p$-value distributions by variant count for true genotypes for original SKAT model versus `parmigiano`-SKAT. Variant bin count is from original SKAT.

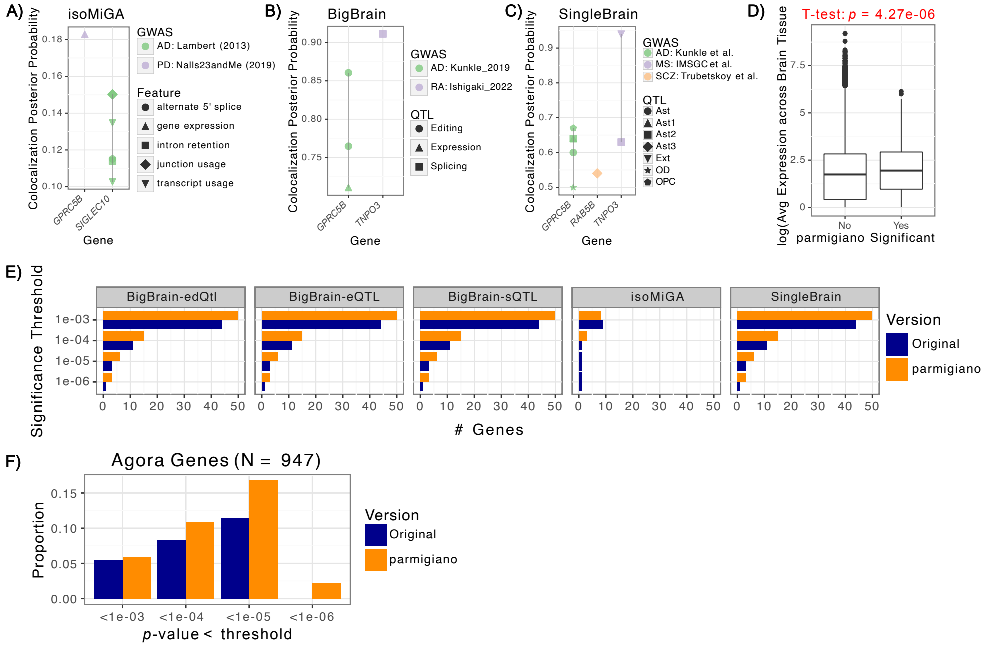


**Fig. S5 Comparison of `parmigiano` results with brain- and AD-relevant datasets. A-C)** Colocalization posterior probabilities for significant discovery genes across **A)** isoMiGA, **B)** BigBrain, and **C)** SingleBrain resources (y-axis), shown per gene (x-axis). **D)** Mean $-\log_{10}(\text{TPM})$ across fourteen GTEx brain tissues for `parmigiano`-significant versus non-significant genes. A one-sided t-test comparing the two groups is denoted in red text. **E)** Number of genes overlapping BigBrain QTLs (editing, expression, splicing), isoMiGA, and SingleBrain QTLs across varying discovery significance thresholds for original and `parmigiano`-integrated tests. **F)** Proportion of genes present in the Agora AD database across discovery significance thresholds for original and `parmigiano` tests.

### Supplemental Tables

**Table S1** Overview of functional annotation groups from WGSA.

| Annotation | Group |
|---|---|
| MAP20 | Conservation |
| phyloP17way_primate | Conservation |
| phyloP30way_mammalian | Conservation |
| phastCons30way_mammalian | Conservation |
| phastCons17way_primate_rankscore | Conservation |
| integrated_fitCons_score | Conservation |
| H1-hESC_fitCons_score | Conservation |
| bStatistic | Conservation |
| GERP_RS | Conservation |
| Roadmap_E074_GenoSkyline_Plus_score | Roadmap |
| Roadmap_E068_GenoSkyline_Plus_score | Roadmap |
| Roadmap_E069_GenoSkyline_Plus_score | Roadmap |
| Roadmap_E072_GenoSkyline_Plus_score | Roadmap |
| Roadmap_E067_GenoSkyline_Plus_score | Roadmap |
| Roadmap_E073_GenoSkyline_Plus_score | Roadmap |
| Roadmap_E070_GenoSkyline_Plus_score | Roadmap |
| Roadmap_E030_GenoSkyline_Plus_score | Roadmap |
| Roadmap_E050_GenoSkyline_Plus_score | Roadmap |
| Roadmap_E051_GenoSkyline_Plus_score | Roadmap |
| Roadmap_E124_GenoSkyline_Plus_score | Roadmap |
| funseq2_noncoding_score | Pathogenicity |
| fathmm-MKL_non-coding_score | Pathogenicity |
| fathmm-MKL_coding_score | Pathogenicity |
| fathmm-XF_score | Pathogenicity |
| CADD_raw | Pathogenicity |
| CADD_phred | Pathogenicity |
| DANN_score | Pathogenicity |
| Eigen-raw | Pathogenicity |
| Eigen-PC-raw | Pathogenicity |
| gnomAD_genomes_POPMAX_AF | gnomAD MAF |
| gnomAD_genomes_AFR_AF | gnomAD MAF |
| gnomAD_genomes_AMR_AF | gnomAD MAF |
| gnomAD_genomes_NFE_AF | gnomAD MAF |
| SpliceAI_DS_AG | Splicing |
| SpliceAI_DS_AL | Splicing |
| SpliceAI_DS_DG | Splicing |
| SpliceAI_DS_DL | Splicing |

### Supplemental Methods

#### 4.9 Functional annotation overview

We integrate variant-level functional annotations from multiple resources to capture diverse biological signals. All annotations are scaled between 0 and 1, where a larger value represents increased predicted variant function.

**Table S2** List of transcription factors for each cell type.

| Cell Type | Transcription Factors |
|---|---|
| Astrocytes | MEIS2, NFIB, TGIF1, EMX2, LHX2, RFX2, RFX4, RORA, RORB, SOX1, SOX2, SOX21, SOX9, NFATC4, SOX5, POU3F2, POU3F3, POU3F4, FOXG1, FOXO1, SP5 |
| Microglia | ETS2, FLI1, SPI1, IRF8, PRDM1, CEBPA, CEBPB, CEBPD, CEBPE, KLF11, KLF2, TFEC, BACH1, BATF, BATF3, RUNX1, RUNX2, RUNX3, LYL1, TAL1, ELK3, MEF2C, CREB3L2, ATF4, MAFB, SALL1, eGFP-SALL1, SMAD5, MEF2A, MEF2B, SMAD2, USF1, STAT3, eGFP-MAFG, NFYB, NRF1, CREB1, IRF1, SOX9 |
| Oligodendrocytes | SOX10, SOX13, SOX21, SOX3, SOX6, SOX8, NHLH2, NFIX, SP7, NFE2, E2F1, CREB5, POU3F3, MYCN |
| Neuron | ASCL1, ASCL5, BHLHA15, BHLHE22, NEUROD1, NEUROD2, NEUROD6, TWIST2, EGR4, SP8, SP9, MEIS3, PKNOX2, MSC, KLF5, KLF8, TBR1, HLF |

***WGSA-derived annotations:***

The majority of the annotations we use are from the Whole Genome Sequencing Annotation database [87]. They are detailed in Table S1. Because many annotations measure related biological processes and exhibit strong correlation structure, we reduce dimensionality using non-negative matrix factorization (NMF), following strategies similar to STAAR [38]. NMF preserves non-negativity and interpretability, allowing annotations to retain directional scaling while capturing shared structure.

Based on correlation patterns and biological interpretation, we define five major functional categories derived from WGSA annotations: splicing, conservation, integrative pathogenicity predictions, brain-specific epigenetic marks from Roadmap [82], and population-specific minor allele frequency (MAF) from gnomAD [83]. For splicing, which consists of four SpliceAI predictions that are sparsely distributed and weakly correlated, we instead use the maximum SpliceAI score per variant rather than applying NMF.

***Cell-type-specific annotations:***

To capture regulatory effects in disease-relevant regions, we include cell-type–specific annotations for microglia, oligodendrocytes, astrocytes, and neurons. Predicted CREs are defined using Activity-by-Contact (ABC) scores [50]. Variants located within predicted CREs are assigned the corresponding ABC score for each cell type.

We further incorporate variant effect predictions from deep learning sequence models. Using Enformer [33], we compute transcription factor (TF) delta scores across 5,318 functional genomics assays. For each variant, we compare model predictions for the reference sequence with those for the alternate allele sequence centered on the variant. Delta scores are defined as the difference between summed predicted signal across the middle 32 bins (4096 bp) of the model output. Predictions are averaged across forward and reverse-complement orientations of the input sequence. To ensure comparability across assays, we normalize scores by computing Z-scores relative to approximately 18 million UK Biobank variants used in PolyFun [88, 89]. Within each cell type, we summarize variant effects using the absolute maximum and absolute minimum delta scores across cell-type–relevant TFs (Table S2).

For ChromBPNet [34], we trained separate models on ATAC-seq data for each of the four brain cell types (astrocyte, microglia, neuron, and oligodendrocyte) using data from Nott et al. [78]. For each variant, scoring was restricted to variants located within a 1,057 bp window (the model context window) of known chromatin accessibility peaks for that cell type; variants falling outside any such window were assigned a score of zero. For each variant–peak pair, we extracted a 2,114 bp genomic sequence centered on the peak and generated both reference and alternate allele sequences. These were scored using an ensemble of five ChromBPNet models per cell type. We quantify allelic effects using the log count difference (lcd) between alternate and reference predictions; when a variant overlapped multiple peaks, the final score was taken as the maximum lcd across peak pairs. These scores quantify predicted regulatory activity and complement TF binding–based signals derived from Enformer.

***Coding context annotations:***

We use binary intron, exon, and UTR (combined 3' and 5') annotations from GENCODE [77]. High confidence LoF and missense predictions are derived from LOFTEE [51]. We additionally incorporate continuous AlphaMissense scores, which predict the pathogenicity of missense variants by estimating the functional impact of amino acid substitutions in protein-coding genes [52]. AlphaMissense scores are defined for variant–gene pairs where the variant falls within a coding region; we restricted scoring to genes expressed in brain-relevant cell types (microglia, astrocytes, excitatory neurons, inhibitory neurons, and oligodendrocytes) using single-cell expression data from the Seattle Alzheimer's Disease Brain Cell Atlas (SEA-AD: https://brain-map.org/consortia/sea-ad).

***Annotation scaling and use in testing***

All continuous annotations undergo PHRED scaling followed by min–max normalization to the range [0,1]. Binary annotations are encoded as 0/1 indicators. This harmonized scaling ensures that annotations contribute comparably when jointly modeled within `parmigiano`. Cell-type–specific tests incorporate both coding and non-coding variants and leverage the full annotation set. Coding-only tests restrict analyses to coding-context annotations, AlphaMissense, conservation, LoF, missense, pathogenicity, splicing, UTR, and population-specific MAF. Coding variants are defined as variants located within exonic regions.
